# Analytical validation of a multi-protein, serum-based assay for disease activity assessments in multiple sclerosis

**DOI:** 10.1101/2022.05.23.22275201

**Authors:** Ferhan Qureshi, Wayne Hu, Louisa Loh, Hemali Patel, Maria DeGuzman, Michael Becich, Fatima Rubio da Costa, Victor Gehman, Fujun Zhang, John Foley, Tanuja Chitnis

## Abstract

**Purpose:** To characterize and analytically validate the MSDA Test, a multi-protein, serum-based biomarker assay developed using Olink^®^ PEA methodology.

**Experimental design:** Two lots of the MSDA Test panel were manufactured and subjected to a comprehensive analytical characterization and validation protocol to detect biomarkers present in the serum of patients with MS. Biomarker concentrations were incorporated into a final algorithm used for calculating four Disease Pathway scores (Immunomodulation, Neuroinflammation, Myelin Biology, and Neuroaxonal Integrity) and an overall Disease Activity score.

**Results:** Analytical characterization demonstrated that the multi-protein panel satisfied the criteria necessary for a fit-for-purpose validation considering the assay’s intended clinical use. This panel met acceptability criteria for 18 biomarkers included in the final algorithm out of 21 biomarkers evaluated. VCAN was omitted based on factors outside of analytical validation; COL4A1 and GH were excluded based on imprecision and diurnal variability, respectively. Performance of the four Disease Pathway and overall Disease Activity scores met the established acceptability criteria.

**Conclusions and clinical relevance:** Analytical validation of this multi-protein, serum-based assay is the first step in establishing its potential utility as a quantitative, minimally invasive, and scalable biomarker panel to enhance the standard of care for patients with MS.

**What is known and what is new in your work?:** *What’s known:*

- Multiple sclerosis (MS) has a complex disease course with variable clinical outcomes; early diagnosis and treatment are critical to management of MS.
- One key focus in MS research is the identification of biomarkers in biological fluids, such as cerebrospinal fluid or blood, to track pathogenesis, disease activity, and disease progression, which may lead to individualized disease management and improved quality of care.
- There currently are no validated clinical tests that leverage multiple blood biomarkers to track disease activity or progression in patients with MS.

*What’s new:*

- The MS Disease Activity (MSDA) Test is a multi-protein, serum-based biomarker assay designed to quantitatively measure disease activity using the protein levels of biomarkers present in the serum of patients with MS.
- In this study, we evaluated 21 biomarkers, 18 of which were selected for inclusion in the MSDA Test, and extensively characterized the MSDA Test (individual biomarkers and algorithmic scores) by establishing the accuracy, precision, sensitivity, and robustness of the assay.
- This study serves as a critical first step in the validation of this multi-protein, serum-based assay, which will be a quantitative, minimally invasive, and scalable tool to improve MS disease management.

**Clinical relevance:** Multiple sclerosis (MS) is a chronic, neurodegenerative, immune-mediated disease of the CNS. MS has a complex disease course with variable clinical outcomes. Although many treatments are effective in early stages of the relapsing/remitting form of the disease, early diagnosis and treatment are critical to managing disease activity and slowing disease progression. One of the major areas of focus in MS research is the identification of biomarkers in biological fluids, such as cerebrospinal fluid or blood, to track pathogenesis, disease activity, and disease progression, which can lead to individualized disease management and improved quality of care. Currently, there are no validated clinical tests that leverage multiple blood biomarkers to track disease activity or progression in patients with MS. Herein, we describe the analytical characterization and validation of a multi-protein, serum-based assay panel developed using Olink^®^ PEA methodology. We demonstrate the extensive characterization of this multi-protein, serum-based assay and establish its accuracy, precision, sensitivity, and robustness. This report will be followed by a complementary clinical validation study investigating the correlation between the proteomic assay results and relevant clinical and radiographic endpoints for patients with MS.

## 1 INTRODUCTION

Multiple sclerosis (MS) is a chronic, neurodegenerative, immune-mediated disease of the CNS, characterized by inflammatory demyelination and neuronal damage.[1,2] MS has a complex disease course with variable symptoms or manifestations that can range from mild and self-limiting to severe.[1] The clinical course, after the first clinical manifestation of the disease, or clinically isolated syndrome, can vary.[3] The damage caused by MS typically leads to relapses, or acute attack of symptoms, followed by progressive disease.[4] Most treatments are effective in early stages of the relapsing/remitting form of the disease;[4,5] however, a delay in treatment can lead to irreversible damage.[6] Studies show that the extent of remyelination in early MS is greater than in chronic MS.[7] Clinical studies are underway to explore treatments targeting remyelination, which may slow or offset disease progression.[8]

The McDonald Criteria, designed to improve the accuracy of MS diagnosis, established the use of MRI to show the accrual of lesions over time and space.[9] The revised McDonald Criteria substituted CSF oligoclonal immunoglobulin G bands for the second clinical/MRI finding.[10] Nonetheless, use of any of these assessments do not always accurately predict disease activity, course, progression, recurrence, or response to treatment.[11-13] As such, there is an unmet clinical need for objective and quantitative measures that can accurately diagnose MS, monitor disease activity, and promote individualized disease management.[13,14]

One major area of focus in MS is the identification of biomarkers in biological fluids, such as CSF or blood, to track pathogenesis, disease activity, and progression.[14,15] One of the key therapeutic strategies in MS is to reduce relapse, lesions, and brain atrophy at all disease stages.[4] As a result, new biomarkers for early MS diagnosis and disease activity monitoring can lead to prevention of disease progression, potentially reducing the patient’s level of disease worsening.[14] The dynamic range of proteins in CSF presents challenges when differentiating small disease-specific changes from inherent inter-individual differences, especially as it relates to methodological variations.[16,17] CSF collection also requires invasive procedures, such as lumbar puncture. On the other hand, blood-based collection of biomarkers allows for safe, quick, and easy collection.[14] With these considerations, detection of biomarkers in blood is a viable and attractive option for the accurate diagnosis and assessment of disease activity and progression in MS. However, there currently are no validated clinical tests that leverage multiple blood biomarkers to track disease activity or progression in patients with MS.[18]

Development of multi-protein assays can be challenging. Each protein biomarker requires specific conditions and methodologies for optimal quantification. The optimal multi-protein assay should be designed so that stability and integrity of all biomarker proteins are maintained and optimized to eliminate cross-reactivity.[19] Larger scale, proteomic techniques allow higher throughput of samples and more timely readout. However, maintaining robustness, repeatability, and sensitivity is challenging, yet critical, to the validation of a multi-protein biomarker panel.[20]

Analysis of multiple proteins may more accurately represent the various pathways, processes, and cell types involved in complex disease states and has the potential to deliver more personalized medicine for MS.[20-23] Single proteins may not perform well alone as diagnostic or prognostic markers. However, as part of a multi-protein assay, they may contribute to a clinically useful model when combined with other proteins and biomarkers.[21] Therefore, multi-protein assay platforms have been characterized and validated for complex disease states.[19,21,22,24]

The MSDA Test is a multi-protein, serum-based biomarker assay designed to quantitatively measure disease activity using the protein levels of biomarkers present in the serum of patients with MS. Our custom multi-protein assay panel was developed using the Olink^®^ PEA (Olink Proteomics, Uppsala, Sweden) methodology described previously (**Figure S1**).[19] Herein, we describe the comprehensive analytical characterization and validation of the MSDA Test to satisfy the criteria necessary for a fit-for-purpose validation considering the assay’s intended clinical use.

## 2 EXPERIMENTAL SECTION

### 2.1 Assay development

Twenty-one biomarkers were selected for inclusion in the custom assay panel based on statistical associations with clinical and radiographic endpoints as demonstrated in feasibility studies for which >1400 proteins were screened using 2 immunoassay platforms **(Table S1)**. These feasibility studies investigated biomarker associations (single-protein and multi-protein) in both cross-sectional and longitudinal samples relative to several radiographic and clinical MS endpoints, including clinically defined relapse versus remission (exacerbation versus quiescence), the presence and count of gadolinium-enhanced lesions on a matched MRI, annualized relapse rate, and Expanded Disability Status Scale. From these studies, the custom panel of 21 proteins was selected with a primary focus on the detection and prediction of disease activity status. The 21 proteins were chosen based on their statistical significance relative to the aforementioned endpoints and with the intent to comprehensively survey the biological pathways, mechanisms, and cell types associated with MS pathophysiology as determined via literature review, protein-protein interaction modeling, gene set enrichment, and spatial expression profiling.[25] Dynamic range of the individual protein assays was considered, as well as the intent to develop a single multi-protein immunoassay panel for which each protein could be measured in an undiluted serum sample. The MSDA Test algorithm consisting of 18 biomarkers included in the panel was finalized in a subsequent clinical validation study for which independent sample sets were analyzed. The final model was trained and validated relative to the presence and count of gadolinium-enhanced lesions.

Serum pools (*n*=4) were included on all runs during assay discovery and development. They were procured in large volumes, aliquoted, stored at −65°C, and run in triplicate. Serum pools were used solely to assess the analytical performance of the assays and served as process controls to determine acceptability of future analytical runs. The SD of repeated measurements was applied to the expected concentrations. Two assay kit lots of the panel were manufactured for which critical reagents were varied to the extent possible.

### 2.2 Description of the two-layer stacked classifier algorithm for determination of the overall Disease Activity score

A two-layer, L2-penalized logistic regression stacked classifier model was developed and clinically validated in a separate study that optimized the model’s performance to classify serum samples based on the presence of gadolinium-enhancing lesions (0 lesions or ≥1 lesions) on an MRI administered within 60 days of blood draw.[26] In the first layer of the model, individual protein concentrations in log_10_ which were demographically corrected for age and sex and LOQ-imputed (referred to as adjusted concentrations) were used as inputs into the four Disease Pathway models (Immunomodulation, Neuroinflammation, Myelin Biology, and Neuroaxonal Integrity). The second layer of the model used the adjusted protein concentrations and the output (eg, the probability) of the Disease Pathway models as meta features to calculate an overall Disease Activity score (**File S1, Supporting Information**). Thresholds were established, which corresponded to low (1.0-4.0), moderate (4.5-7.0), and high (7.5-10.0) Disease Activity scores. Analytical characterization and validation of the individual biomarkers were factors used to determine inclusion of those biomarkers in the algorithm.

### 2.3 Incurred sample reanalysis

Incurred sample reanalysis was performed to characterize precision and robustness for the individual biomarkers and the Disease Activity and Disease Pathway scores. Forty-eight individual samples from patients with MS were repeatedly analyzed across 10 plates over ≥5 days with varied equipment, reagents, location, and personnel. Acceptability criteria for individual biomarkers was an average %CV ≤20%, and average SD at all established Disease Activity score levels of ≤1.0 units. The 48 samples broadly represented the expected range of biomarker values and Disease Activity scores in the real-world MS population.

### 2.4 Assay accuracy, precision, and sensitivity

Accuracy for each analyte was determined by mixing serum samples at different ratios and evaluating the percent recovery of the observed concentration relative to the expected concentration. Sample mixing enabled the accuracy assessment to be performed using endogenous protein versus a recombinant protein source. Expected concentrations were calculated by applying the targeted ratios of unmixed samples. The ratios of sample mixtures with two samples were 25%:75%, 50%:50%, and 75%:25%. The ratios of sample blends for mixtures with four samples were 25%:25%:25%:25% and 40%:10%:40%:10%. Additionally, accuracy was also evaluated for the Disease Pathway and Disease Activity algorithms by correlating observed scores with expected scores using the same sample mixtures created for the individual analyte assessments.

Intra- and inter-assay precision was measured for each analyte. The %CV was determined using serum pools enabling the assessment to be performed using endogenous protein. Serum pools were manufactured to represent patients with shorter and longer MS disease duration, those with inflammatory disease (rheumatoid arthritis), and one healthy control. Acceptability criteria for intra- and inter-assay precision was established as %CV ≤15% and ≤20%, respectively.

Sensitivity was defined as the assay’s ability to accurately and precisely detect low concentrations of a given substance in biological specimens. To establish the ULOQ and LLOQ, a LOQ panel was manufactured during assay development. For each analyte, four levels were targeted near the anticipated upper limit (ULOQ 1-4) and four levels were targeted near the anticipated lower limit (LLOQ 5-8). The targeted concentrations were based on expected real-world MS patient sample distributions, the shape of the standard curve, and location of asymptotes. The LOQ panel was run in triplicate over two lots (≥5 runs per lot) and fit to the standard curve. Accuracy, defined as 80-120% recovery relative to the expected concentration and precision (inter-assay %CV ≤20%), were used to establish the acceptability criteria and determine the LLOQ and ULOQ of each analyte. Additionally, individual LOQs were assessed and established separately for each kit lot. The most conservative LOQ levels with acceptable accuracy and precision parameters for both lots were used to establish the final LLOQ and ULOQ.

Undiluted serum samples were run in the MSDA Test and as a result, no dilution factor was accounted for in the sensitivity analysis. Therefore, the LLOQ and ULOQ define both the analytical measurement range and the reportable range of the assay. Serum samples that recovered either above the ULOQ or below the LLOQ were reported at the established LOQ concentration (referred to as LOQ imputation). MS serum samples were used to establish MS reference ranges for each biomarker. A diverse set of patient samples were used throughout the assay development process and for the analytical validation studies. A total of 1645 samples from nine deeply phenotyped cohorts were analyzed primarily for evaluating associations of biomarkers with MS disease activity and disease progression endpoints. Additional samples from both patients with MS and other disease states were procured for specific analytical characterization experiments. The 1645 samples that were analyzed for the associations of biomarkers with MS endpoints were combined in the subsequent analysis to establish MS reference ranges. These samples were collected both retrospectively and prospectively from nine US and international sites and broadly represent the real-world MS population. The mean ± SD age of these patients at the time of the blood draw was 40.85 ± 11.0 years, with a mean ± SD disease duration of 8.39 ± 8.0 years; 72.8% of the patients were female. For race, the top 3 categories were White (81.4%), unknown/not reported (13.5%), and Black/African American (2.7%). The primary endpoint used to train the finalized MSDA Test algorithm was the presence and count of gadolinium-enhancing lesions on an MRI administered within close proximity to the blood draw. For the 1645 patient samples, 1326 had available gadolinium-positive (Gd+) lesion counts and 53.0% of the patient samples had ≥1 Gd+ lesion. The linear interpolation method was used to establish the 95% interval (2.5th and 97.5th percentiles).[27] The percentile relative to these reference ranges are presented with their protein concentrations.

### 2.5 Assay interference

Assay interference was defined as the effect of a substance present in the sample altering the correct value of the result or the recovery of samples in the assay. Since patients with MS may be treated with a variety of drugs, potential interference of drugs was tested to determine if their presence would affect measurement of the individual protein biomarkers. Concentrations of common prescriptions, over-the-counter drugs, common MS drugs, and DMTs were spiked into serum samples (**Table S2**). Concentrations of common prescription and over-the-counter drugs were determined by Sun Diagnostics (New Gloucester, ME, USA) using a commercially available test kit. DMTs were targeted at two times C_max_ from pharmacokinetic studies, or the highest possible concentration allowable for spiking with the procured interferent stock. Finally, a universal mAb standard was tested at two concentrations (424 and 7.93 μg/mL) to cover the two times C_max_ of several mAb DMTs. Endogenous substances (hemoglobin, bilirubin, and lipids) and heterophilic antibodies (RF and HAMA) were also measured. For most interferent substances, the acceptability threshold, or median recovery, for the interference assessment was established as 80-120% relative to a corresponding spike control, except for HAMA for which percent recovery of sample mixtures was evaluated (**File S2, Supporting Information**).

### 2.6 Diurnal variability

Patient serum samples were collected at days 1-5 and day 12 to characterize biomarker level fluctuations. For each of the six time points per patient, the %CV and the percentage difference of the observed protein concentration relative to the average concentration at all time points were calculated.

### 2.7 Sample stability

In an initial experiment, stability studies for four serum samples were performed to determine the effect that storage and processing conditions can have in a clinical setting. Stability was assessed at the following four temperatures: −65°C or below (−80°C), −10°C or below (−20°C), 2-8°C (4°C), and room temperature (18-25°C) at the following time points: 4 hours (for 4°C and room temperature) and days 1, 3, 7, 14, and 28 (for −20°C, 4°C, and room temperature). The results from −20°C, 4°C, and room temperature were compared with the control storage condition (−80°C). In a follow-up study, the stability of storage of 14 samples was evaluated at 4°C at days 1-3 and 7 compared with a control storage condition (−80°C) to establish the duration of time samples that can be stored at 4°C. Five freeze-thaw cycles, performed at −65°C or below, were also evaluated using four MS serum samples compared with fresh samples.

## 3 RESULTS AND DISCUSSION

### 3.1 Analytical characterization and validation

Experiments were performed between July 2020 and July 2021. Fifty-one plates were run (40 and 11 plates using the first and second lots of manufactured kits, respectively).

Based on the analytical validation and characterization of individual biomarkers described below, the 18 out of 21 biomarkers that were included in the algorithm were determined to have acceptable analytical performance. GH and COL4A1 were excluded from the algorithm based on the analytical characterization studies described below. VCAN was not incorporated into the final algorithm due to biostatistical factors unrelated to analytical performance.

### 3.2 Incurred sample reanalysis

All individual biomarkers were determined to have a mean %CV <20% and met established acceptability criteria (**Figure 1A**). The Disease Activity score and the four Disease Pathway scores demonstrated reproducible results throughout the range of scores (**Figures 1B-1F)**. For the Disease Activity score, the average SD across 48 samples was observed to be 0.3 score units, which is less than one interval (0.5) on the reportable scale, and as a result, met acceptability criteria. Additionally, incurred sample reanalysis showed robustness and equivalency of the assay between lots and laboratories, with the exception of COL4A1 (**Table S3**).

**FIGURE 1.**
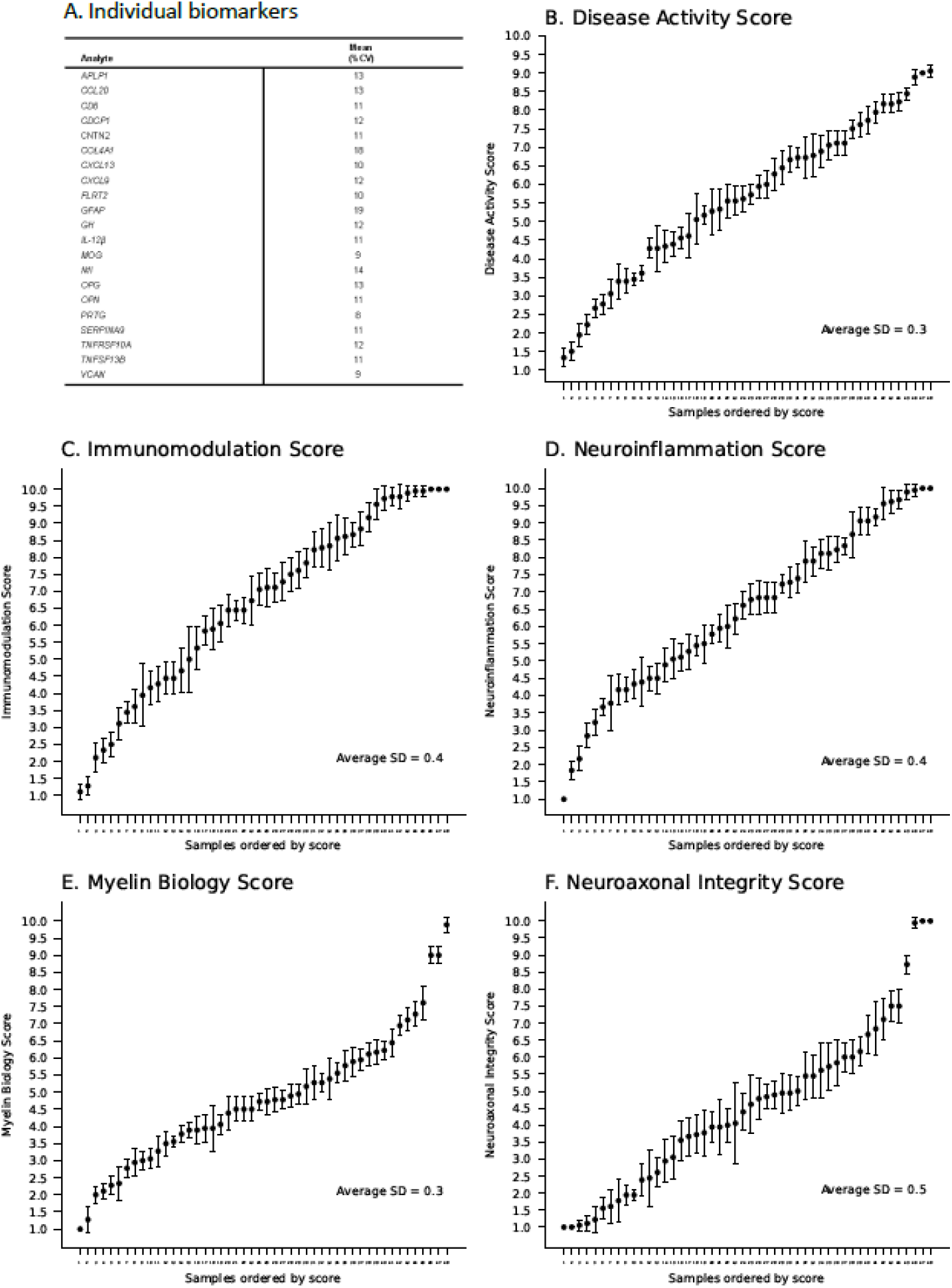
Incurred sample reanalysis results for (A) Individual biomarkers and (B) Overall Disease Activity score, (C) Immunomodulation, (D) Neuroinflammation, (E) Myelin Biology, and (F) Neuroaxonal Integrity pathway scores in the MSDA Test.

### 3.3 Assay accuracy, precision, and sensitivity

Samples for the accuracy assessment were selected from an internal MS cohort (*n*=64) to target both high and low concentrations for the individual biomarkers relative to the MS population. Twenty mixed samples from four selected samples were analyzed for each biomarker. Minimum percent recovery for each biomarker ranged from 78% to 89%; the maximum percent recovery for each biomarker ranged from 99% to 124%. The median percent recovery ranged from 91% to 100% (**Figure 2A**). Additionally, the Disease Pathway and overall Disease Activity scores were calculated for both observed and expected concentrations of the various sample mixtures. The observed calculated scores correlated with the expected scores; R^2^≥0.85 was established as the acceptability criteria (**Figures 2B-2F**).

**FIGURE 2.**
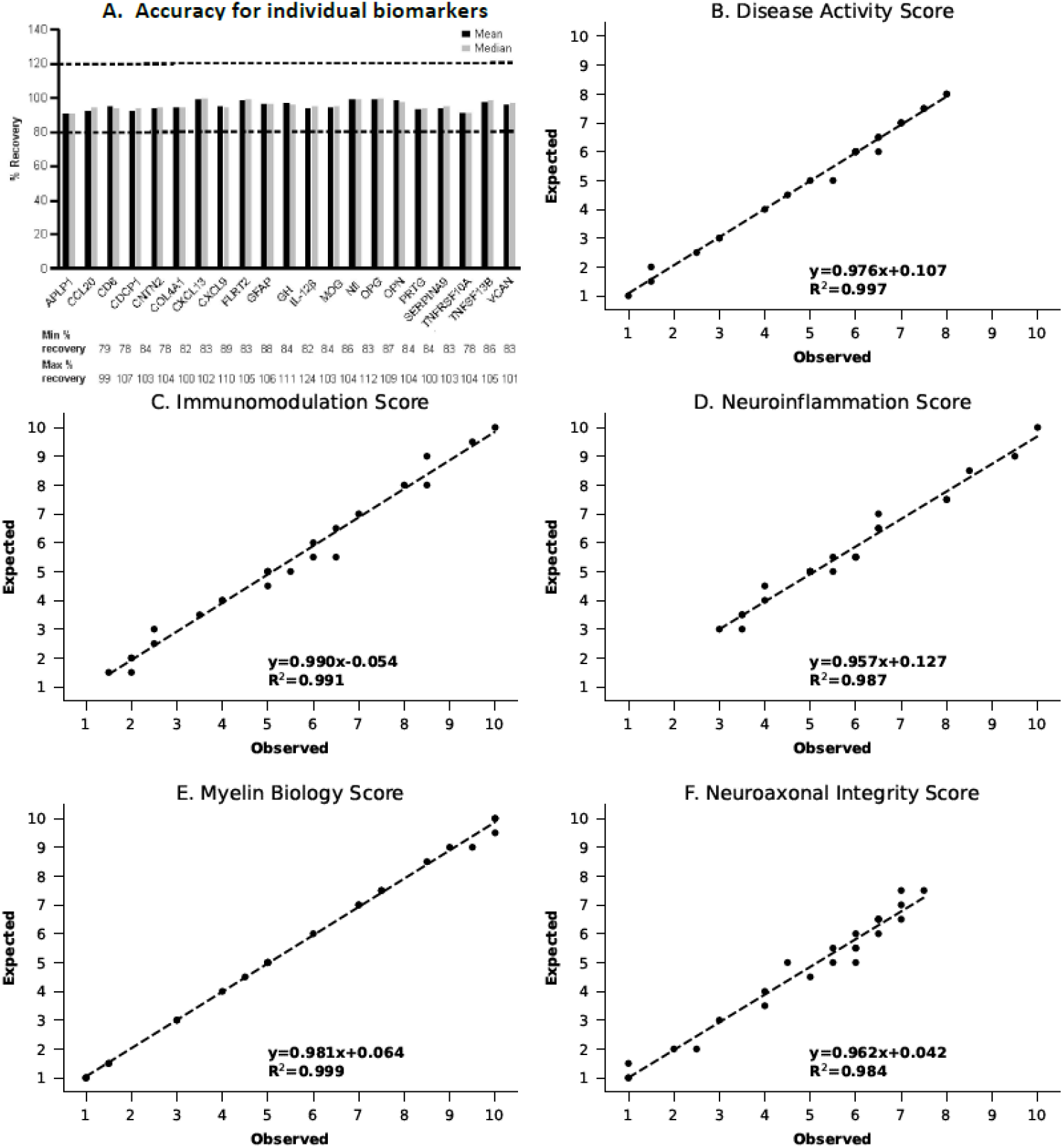
Accuracy of the MSDA Test to detect (A) Individual biomarkers and (B) Overall Disease Activity score, (C) Immunomodulation, (D) Neuroinflammation, (E) Myelin Biology, and (F) Neuroaxonal Integrity pathway scores.

Twelve replicates per serum pool were analyzed on a single plate for the intra-assay precision assessment; ≤51 values per serum pool were analyzed across 51 plates spanning 2 lots of reagent kits. The intra- and inter-assay precision satisfied the criteria for meeting the precision parameter with most analytes passing the established criteria. Of note, COL4A1 was found to have inferior inter- and intra-assay precision that ranged from 7% to 47% and 15% to 59%, respectively. Based on these findings, COL4A1 was removed from consideration for inclusion in the algorithm. MS serum samples (*N*=1645) were analyzed during the assay development and validation process and used to establish the MS reference ranges for each analyte. Sensitivity analysis demonstrated that the LLOQ and ULOQ of each analyte met the sensitivity requirements established for the assay. The maximum percentage of samples requiring imputation at any LOQ was 1.8% (for NfL at LLOQ) (**Table 1**).

**TABLE 1.**
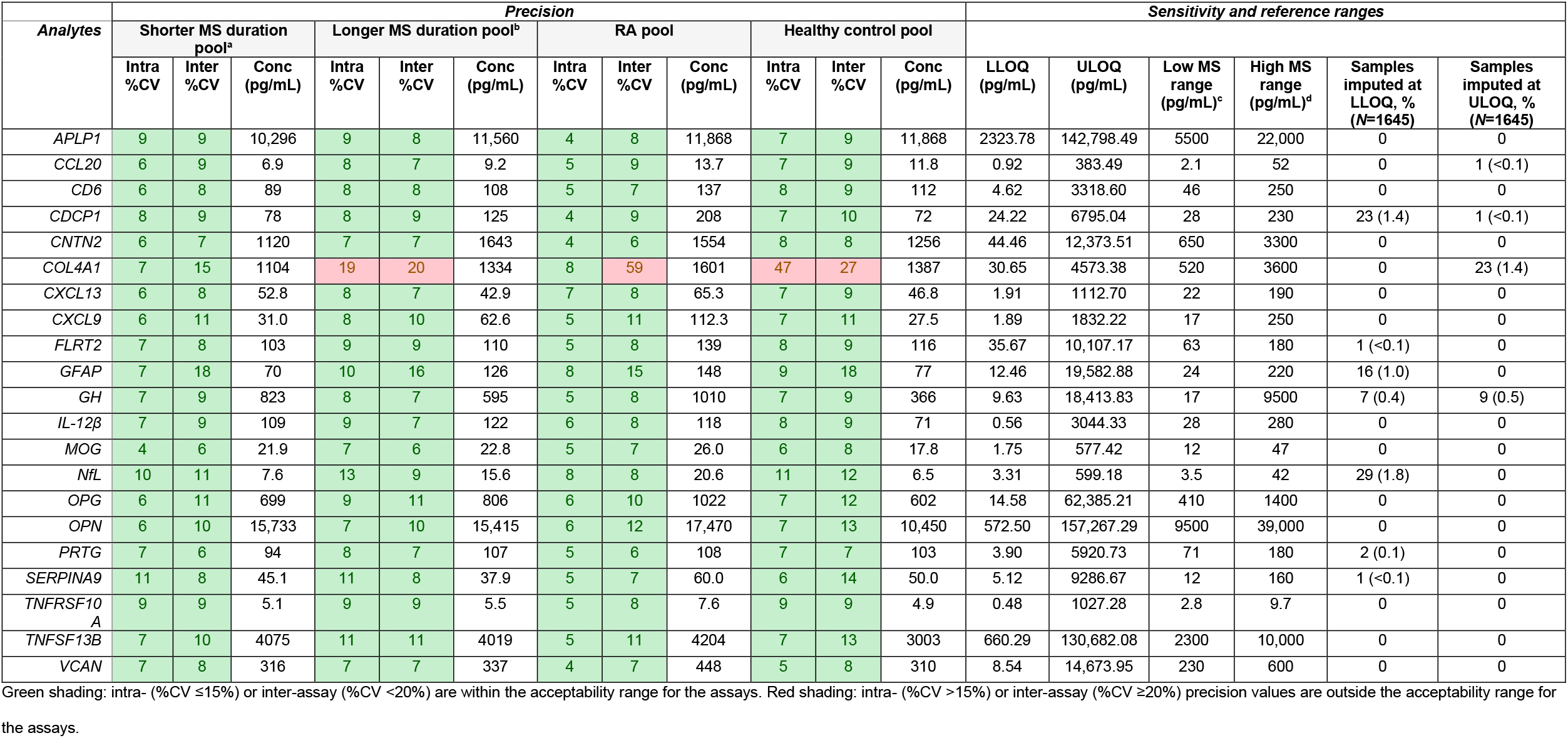
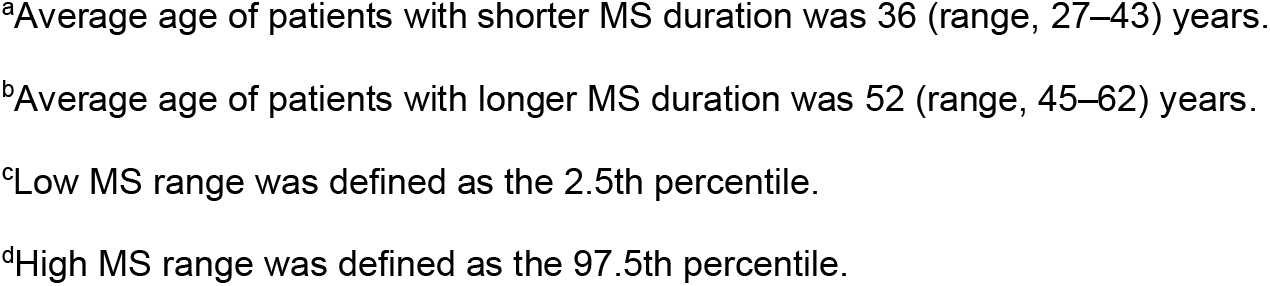
Intra- and inter-assay precision, sensitivity, and reference ranges for biomarkers in the MSDA Test

### 3.3 Assay interference

Most biomarker interactions with interferent combinations, such as common MS drugs, DMTs, and mAbs produced a median recovery that ranged from 80% to 120% (**Figure 3**). A lower percentage recovery was observed for two biomarkers, COL4A1 and CCL20, demonstrating a potential alteration in the presence of the sample for individual drugs. COL4A1 produced a low percent recovery for several drugs that ranged from 71% to 79%, which was likely an artifact of established assay imprecision (**Figure 3** and **Figure S2**). For CCL20, cefoxitin spiked at 660 mg/dL resulted in a median percent recovery of 77% (**Figure S2**). Additional assay interferents are shown in **Figures S2** (common drugs) and **S3** (routine endogenous interferents and heterophilic antibodies).

**FIGURE 3.**
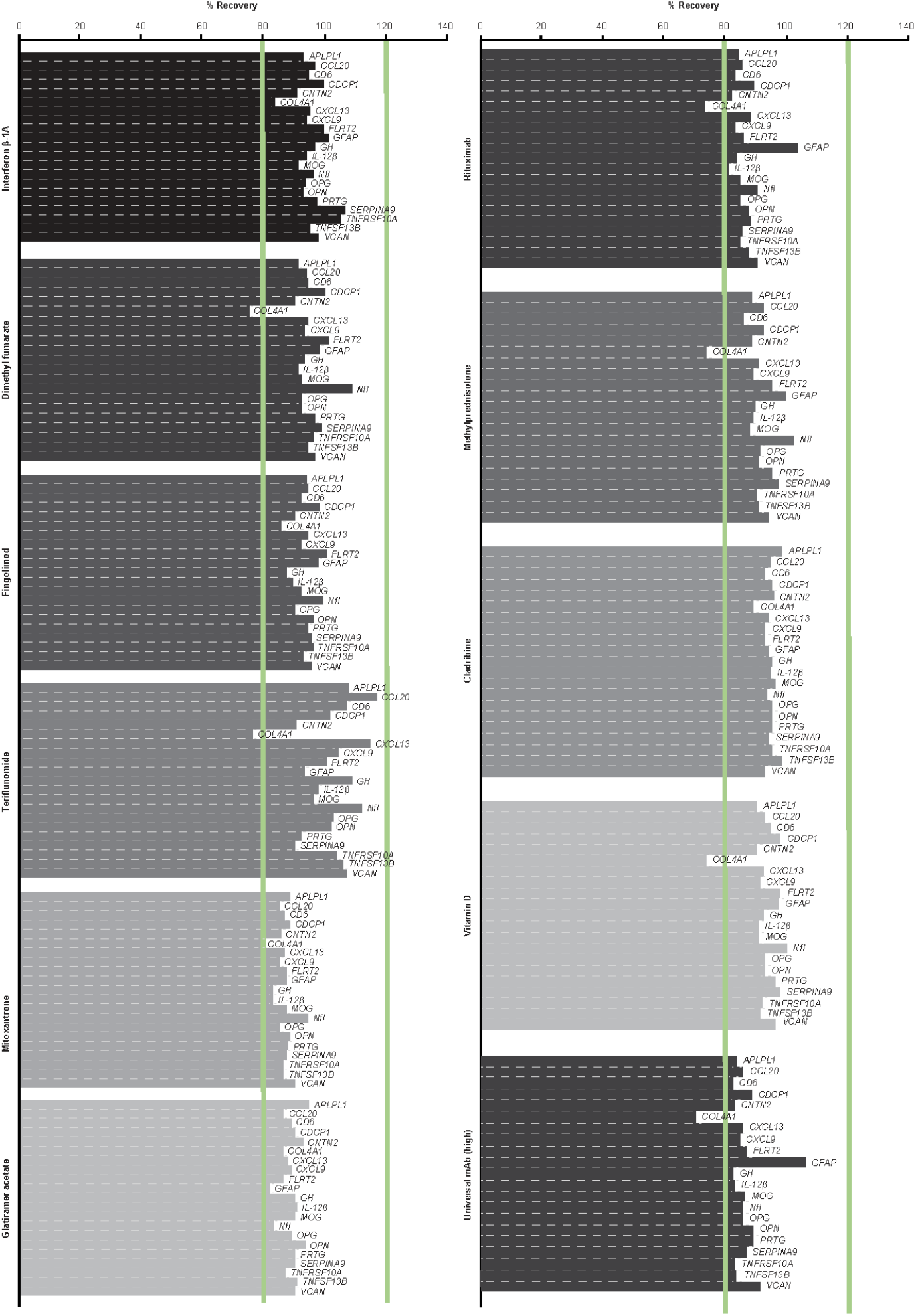
Assay interference for common MS drugs, DMTs, and the high concentration of universal mAb surrogates in the MSDA Test.

### 3.4 Diurnal variability

Diurnal variation was evaluated in eight patients over six time points (**Figure S4**). Mean and median percent differences for each biomarker and patient were observed to be within ±20%; mean and median %CV was found to be <30% for 19 of the 21 biomarkers (**Table 2**). Of note, there were some individual samples that were outside of the acceptable range (±30%; data not shown). In addition, mean and median diurnal variability ≥30% was observed for COL4A1, which may have been due to the imprecision of the assay to detect this biomarker. GH was also found to be more variable compared with the other biomarkers, which is not surprising, as GH has been previously reported to have a high degree of ultradian and diurnal variability.[28] For this reason, GH was removed from consideration for inclusion in the algorithm.

**TABLE 2.** Diurnal variability of eight samples in the MSDA Test across six time points (days 1, 2, 3, 4, 5, and 12)

| ANALYTE | MEAN %CV | MEDIAN %CV |
| --- | --- | --- |
| *APLP1* | 14 | 15 |
| *CCL20* | 25 | 21 |
| *CD6* | 10 | 7 |
| *CDCP1* | 12 | 12 |
| *CNTN2* | 11 | 10 |
| *COL4A1* | 44 | 39 |
| *CXCL13* | 18 | 11 |
| *CXCL9* | 13 | 11 |
| *FLRT2* | 10 | 8 |
| *GFAP* | 13 | 12 |
| *GH* | 78 | 79 |
| *IL-12B* | 10 | 9 |
| *MOG* | 12 | 11 |
| *NFL* | 17 | 17 |
| *OPG* | 11 | 10 |
| *OPN* | 10 | 8 |
| *PRTG* | 8 | 6 |
| *SERPINA9* | 12 | 12 |
| *TNFRSF10A* | 10 | 12 |
| *TNFSF13B* | 11 | 8 |
| *VCAN* | 10 | 7 |

Green shading: %CV ≤30%. Red shading: %CV >30%.

### 3.5 Sample stability

In the initial stability study, all biomarkers were stable for up to 1 day at room temperature and at 4°C, and for 28 days at −20°C. For those samples stored at room temperature, CXCL13, IL-12β, and TNFSF13B decreased beyond −20% at 3 days. During a follow-up study, all biomarkers were found to meet acceptability criteria when stored at 4°C, and consistent with the initial study as well as the control condition (−80°C) at follow-up (**Table S4**). In a study to examine the stability of samples after freeze-thaw, most biomarkers met acceptability criteria when compared with fresh sample. Of note, GFAP concentrations decreased beyond −20% for freeze-thaw cycles 4 and 5 (**Table S5**). Finally, score level analysis showed that test conditions were within 3 SDs (±1.5 score difference) from the control conditions during the initial study (**Table S6**) and at follow-up (**Table S7**). From these findings, we showed that biomarker levels were found to be most affected above certain thresholds (room temperature for 24 hours, 4°C for 7 days, −20°C for 28 days, and three freeze-thaws). These data can be used to establish allowable sample handling and storage conditions. Beyond these empirical estimations of protein stability, it is also important to note that statistically meaningful associations of biomarkers with multiple MS endpoints were observed using samples that had been stored at −80°C for extended periods of time. This suggests that the target epitopes for the proteins that were selected for inclusion in the custom assay panel and the final algorithm were sufficiently stable to derive clinically meaningful insights.

## 4 CONCLUSION

The accuracy, precision, and reproducibility of a biomarker assay are critical to its utility as a diagnostic and prognostic tool in the management of complex neurodegenerative disorders such as MS. Additionally, such an assay should be insensitive to external factors such as assay interferents and sample collection, processing, and storage. The Clinical and Laboratory Standards Institute and the United States Food and Drug Administration issued guidance on the development and validation of assays for the detection of serum-based biomarkers.[27,29,30] Parameters such as accuracy, precision, recovery, sensitivity and specificity, quality control, and sample stability need to be optimized for the assay to be properly validated.[27,29,30] Results from our analytical validation experiments to characterize the MSDA Test support that the assay is accurate, precise, sensitive, specific, and robust at determining individual biomarker levels and algorithmic scores, regardless of assay interferents, and validated in terms of sample stability. Our findings of high accuracy and precision for the MSDA Test assay align with those of other validation studies of multi-protein assays utilizing the same,[31,32] as well as alternative[21,22] platforms.

PEA demonstrated high sensitivity, specificity, reproducibility, and repeatability with low intra- and inter-assay variability, which has allowed for large-scale, high throughput screening of up to 92 proteins in 96 samples simultaneously, with low sample consumption and cost.[19] This platform detected novel protein biomarkers and biomarker combinations for many complex disease states, such as cardiovascular disease,[33-37] cancer,[32,38-40] Alzheimer’s disease,[41] and inflammatory diseases such as atopic dermatitis and lupus[42,43]; the platform has also proven useful in aging research.[44] For the MSDA Test, we demonstrated that a focused panel of MS biomarkers can be developed and optimized on the PEA platform with absolute quantitation of the proteins to support a fit-for-purpose analytical validation, thereby enabling clinical use of the assay.

Thus far, there are no validated clinical tests that leverage multiple blood biomarkers to track disease activity or progression in patients with MS. This is critical for a disease such as MS, which has a complicated clinical course varying from mild, self-limiting to severe.[1] Although MS disease prognosis is primarily based on clinical evidence, such as relapse rate and disability progression, and diagnostic tests (eg, brain MRI or the presence of oligoclonal immunoglobulin G bands in the CSF),[14] neither can consistently and accurately predict disease course, activity, or prognosis.[13] Given the emphasis on early diagnosis and the efficacy of therapies to treat early stages of the relapsing/remitting form of the disease,[4,5] validation of a biomarker panel remains an unmet need in clinical practice, and use of this biomarker tool should provide diagnostic and prognostic value for the treatment of MS. This study demonstrated identification of biomarkers for this complex disease using the PEA platform. With further clinical validation, this assay can potentially be used to track disease activity and progression of MS, allowing a more personalized approach to MS treatment.

A limitation of using a multi-protein assay is that the conditions established for one biomarker are not always uniform across the full panel of biomarkers. Our findings show that the MSDA Test was optimized for assessment of 18 out of the 21 included biomarkers and the analytical validation paradigm that we described demonstrates a high level of accuracy, sensitivity, and precision with minimal cross-reactivity and interference by substances commonly seen in patients with MS.

This study serves as a critical first step in the validation of a multi-protein, serum-based assay. The next step in the validation of the MSDA Test is clinical validation, which will support and confirm the association between the serum-based MSDA Test and clinical and radiographic MS endpoints. Upon completion of clinical validation of the assay, the final Disease Activity and Disease Pathway algorithms will use the ensemble of validated proteins to expand the use of the assay by evaluating biomarker correlations with endpoints associated with additional MS disease assessments, selection of therapy, and differential diagnosis of patients with MS. Upon successful clinical validation, this MSDA Test will be a quantitative, minimally invasive, and scalable tool to improve disease management for patients with MS and their physicians.

## Data Availability

Access to data can be provided after a research proposal is submitted to the corresponding author and a data sharing agreement is in place.

## Abbreviations

%CV: percent coefficient of variation
APLP1: amyloid beta precursor-like protein 1
CCL20: C-C motif chemokine ligand 20
CD6: cluster of differentiation 6
CDCP1: CUB domain-containing protein 1
C_max_: maximum concentration
CNS: central nervous system, CNTN2, contactin 2
COL4A1: collagen type IV alpha-1
conc: concentration
CSF: cerebrospinal fluid
CXCL9: chemokine (C-X-C motif) ligand 9 (MIG)
CXCL13: chemokine (C-X-C motif) ligand 13
DMT: disease-modifying therapy
DNA: deoxyribonucleic acid
FLRT2: fibronectin leucine-rich repeat transmembrane protein
Gd+: gadolinium positive
GFAP: glial fibrillary acidic protein
GH: growth hormone
HAMA: human anti-mouse antibodies
HCl: hydrochloride
IL-12β: interleukin-12 subunit beta
LLOQ: lower limit of quantitation
LOQ: limit of quantitation
mAb: monoclonal antibody
Max: maximum
Min: minimum
MOG: myelin oligodendrocyte glycoprotein
MRI: magnetic resonance imaging
MS: multiple sclerosis
MSDA: Multiple Sclerosis Disease Activity
Na: sodium
NfL: neurofilament light chain
OPG: osteoprotegerin
OPN: osteopontin
PCR: polymerase chain reaction
PEA: Proximity Extension Assay
PRTG: protogenin
qPCR: quantitative polymerase chain reaction
R^2^: coefficient of determination
RA: rheumatoid arthritis
RF: rheumatoid factor
SD: standard deviation
SERPINA9: serpin family A member 9
TNFRSF10A: tumor necrosis factor receptor superfamily member 10A (TRAIL-R1)
TNFSF13B: tumor necrosis factor superfamily member 13B (BAFF)
ULOQ: upper limit of quantitation
VCAN: versican core protein

## ACKNOWLEDGMENTS

The authors wish to thank the following team members from Olink Proteomics (Uppsala, Sweden) who were involved in the development of the Multiple Sclerosis Disease Activity Test assay: Erika Assarsson, Sandra Ohlsson, Martin Lundberg, Jessica Bergman, and Niklas Nordberg. All authors contributed to and approved the manuscript for submission. Writing and editorial assistance were provided by Jennifer L. Venzie, PhD, and Bu Reinen, PhD, CMPP, of The Lockwood Group (Stamford, CT, USA), and were funded by Octave Bioscience, Inc.

## CONFLICT OF INTEREST STATEMENT

F. Qureshi, W. Hu, L. Loh, H. Patel and M. DeGuzman are employees of Octave Bioscience, Inc. M. Becich, F. Rubio da Costa, V. Gehman, and F. Zhang were employees of Octave Bioscience, Inc. at the time the study was completed. J. Foley has received research support from Biogen, Novartis, Adamas, Octave Bioscience, Inc., Genentech, and Mallinckrodt, received speakers’ honoraria and acted as a consultant for EMD Serono, Genzyme, Novartis, Biogen, and Genentech, and has equity interest in Octave Bioscience Inc., and is the founder of InterPro Biosciences. T. Chitnis has received compensation for consulting from Biogen, Novartis Pharmaceuticals, Roche Genentech, and Sanofi Genzyme, and received research support from the National Institutes of Health, National MS Society, US Department of Defense, EMD Serono, I-Mab Biopharma, Mallinckrodt ARD, Novartis Pharmaceuticals, Octave Bioscience, Inc., Roche Genentech, and Tiziana Life Sciences.

## FUNDING

The study was funded by Octave Bioscience, Inc. and in part by the U.S. Department of Defense (W81XWH2110633 to T Chitnis).

## Supporting Information

### FILE S1. MSDA Test algorithm formula

Key to the MSDA Test was the architecture of a stacked classifier (in the mlxtend [1] framework) where each node (blue boxes in the **Figure**) in both layers are L2-regularized logistic regression models from Scikit Learn.[2] The input layer read in demographically corrected for age and sex, log_10,_ LOQ-imputed concentrations of proteins associated with four physiological pathways from each sample and generated probabilities that the sample in question was positive for the presence of Gd+lesions. The meta-classifier (output) layer of the model ingested the probabilities from each pathway model along with all of the protein concentrations that fed the first layer and generated an overall probability that the sample in question was Gd+ lesion-positive. This probability was then mapped into a Disease Activity score by being shifted, scaled, rounded, and clipped to the nearest half point over a range from 1 to 10 (Eq. 1). A similar process converted the pathway probabilities into pathway scores after the pathway probabilities were adjusted so that for each sample, the mean over the adjusted pathway probabilities was equal to the Disease Activity probability for each sample (Eq. 2). The datasets used to develop and evaluate the demographic correction, four Disease Pathway models, and Disease Activity model, as well as the performance of these models, have been described previously.[2]

The process for calculating the Disease Activity score is shown in the **Figure**.

**FIGURE.**
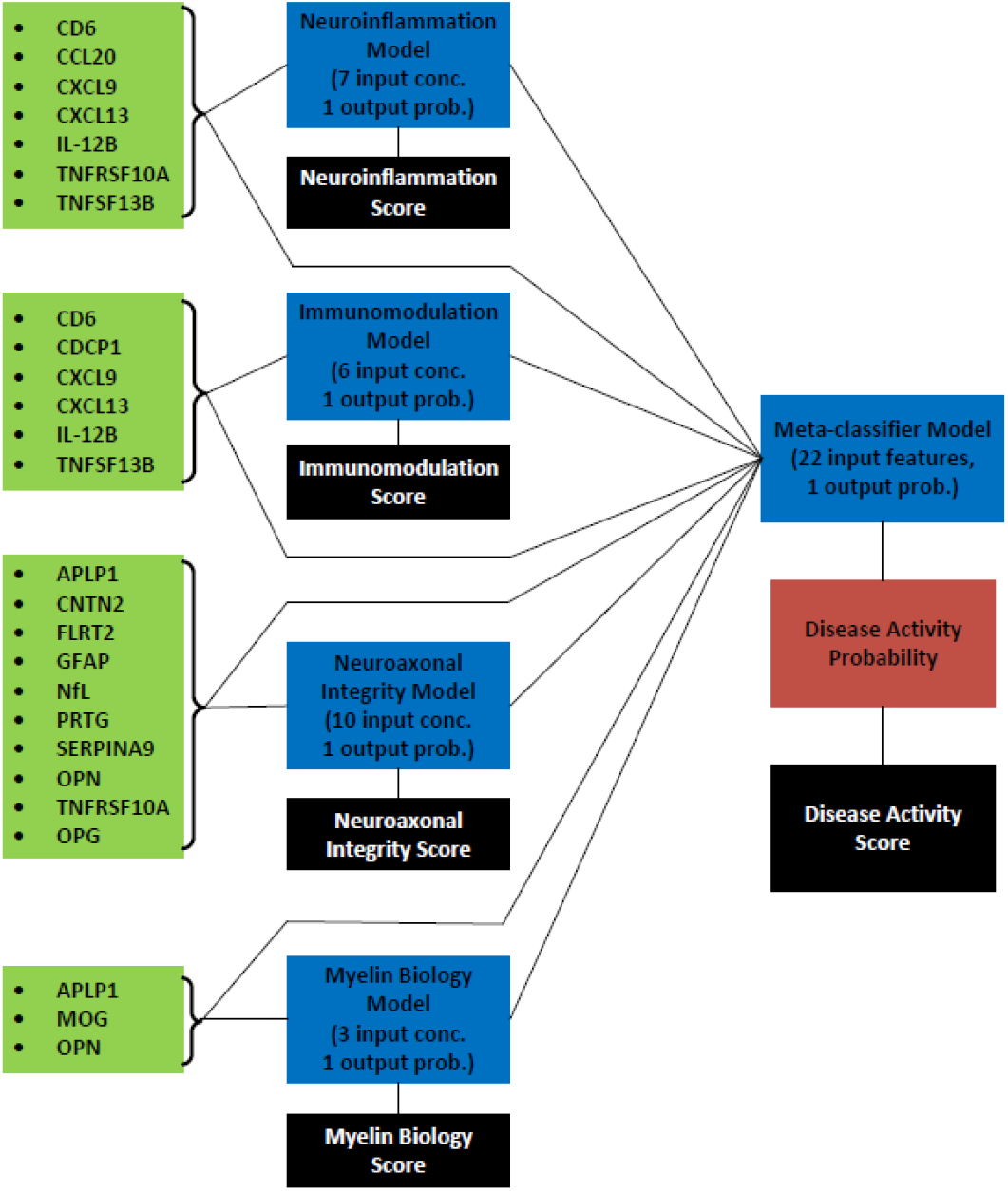
Schematic representation of the Disease Activity score calculation from protein concentrations (green) to both layers of the stacked classifier (blue), the output Disease Activity probability (red), and the final Disease Activity and Disease Pathway score (black).

The overall Disease Activity score had the mathematical form:

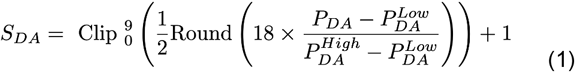

The “Clip” function limited the output of its argument to between 0 and 9. The “Round” function rounded to the nearest whole number. *P*_*DA*_ was the Disease Activity probability output from the final stage of the stacked classifier model. *P*^*High*^_*DA*_ and *P*^*Low*^_*DA*_ were probability clip values chosen for the overall Disease Activity (and each pathway) to maximize the range of the score over the entire training data set. Their values are tabulated below.

Similar to the Disease Activity score, each pathway score was computed from the pathway probabilities with the additional step of recentering the ensemble of pathway probabilities around the Disease Activity probability for each sample:

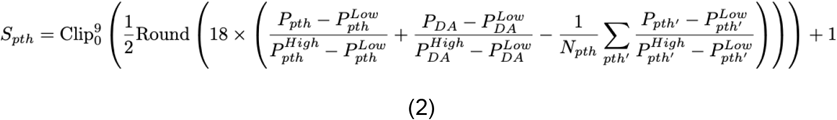

“Clip” and “Round” served the same purpose in Eq. 2 as in Eq.1. *P*_*pth*_ was the probability for pathway model *pth. P*_*DA*_ was again the probability output by the final stage of the stacked classifier. The “High” and “Low” *P*s for each *pth* served a similar purpose to those for the Disease Activity: rescaling the model probabilities so that they had a larger dynamic range. Lastly, *N*_*pth*_ was the number of pathway models that fed into the final layer of the stacked classifier model. In this case, *N*_*pth*_=4.

The probabilities associated with each layer of the stacked classifier had a closed analytic form as well. Working backward from the final Disease Activity score, the Disease Activity probability had the form:

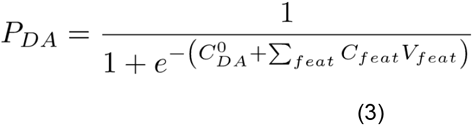

*C*^*0*^_*DA*_ was the intercept (or bias) value for the Disease Activity meta-classifier. *C*_*feat*_ was the coefficient associated with *V*_*feat*_. *V*_*feat*_ was the value of the feature being summed, either the probability for one of the pathways from the first layer of the model (written out explicitly in Eq. 4) or the age- and sex-corrected, log_10_, LOQ-imputed concentrations that fed into the first-layer pathway models. These meta-classifier coefficients were fit using L2-penalized logistic regression with Gd+ lesion presence as the dependent variable, an intercept, and the stacked classifier features as the independent variables, with inverse regularization strength C=1.0, balanced class weight, and tolerance =0.0001, with the ‘lbfgs’ solver.

The individual pathway probabilities had the form:

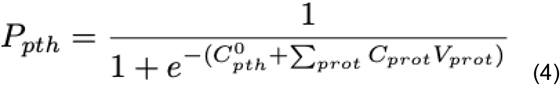

*C*^*0*^_*pth*_ was the intercept value for the pathway model in question. *C*_*prot*_ was the coefficient for a protein in pathway *pth. V*_*prot*_ was the demographically corrected, log_10_, LOQ-imputed concentration of protein *prot*. These coefficients were fit using L2-penalized logistic regression with Gd+ lesion presence as the dependent variable, an intercept, and the pathway’s demographically corrected (for age, sex), log_10_, LOQ-imputed protein concentrations as the independent variables. The final pathway coefficients were the result of a stratified, 10-fold cross-validation with 3 repeats, grid-searched over inverse regularization strength C=[100, 10, 1.0, 0.1, 0.01], class weights=[None, balanced], solvers=[“saga,” “lbfgs,” “liblinear”], and tolerances=[0.01, 0.001, 0.0001]. The demographic correction for the log_10_ of the LOQ-imputed protein concentration had the form:

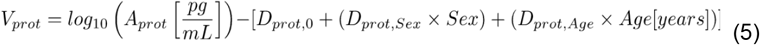

*A*_*prot*_ was the LOQ-imputed absolute concentration of protein *prot* in pg/mL (the units of these direct measurements have been written explicitly for added clarity). *D*_*prot, 0*_, *D*_*prot, Sex*_, and *D*_*prot, Age*_ were the intercept, sex, and age coefficients for demographic correction, respectively. These coefficients were fit using the following procedure on three datasets not reported here. The dataset used to train the stacked classifier and pathway models provided the final non-zero coefficients for the demographic correction. The demographic correction coefficients were fit independently on each dataset using linear regression using StatsModels OLS[4] with log_10_, LOQ-imputed protein concentration as the dependent variable, an intercept, and age and sex as the independent variables, after outliers were removed. Outliers were defined for each dataset as ≥96th percentile or ≤4th percentile for log_10_, LOQ-imputed protein concentration. After fitting the linear regression, the age and sex coefficients were set to zero if the 95% confidence interval contained zero in any of the three datasets or if the sign of the coefficient was not consistent across three datasets. Sex was 0 for women and 1 for men, and age was the patient’s age in years at time of blood draw.

Finally, see below for tables of the coefficients used in each step of the scoring process. First, we tabulated the demographic correction coefficients. These were the *D*s in Eq. 5.

| Protein | Sex | Age | Intercept |
| --- | --- | --- | --- |
| APLP1 | -0.052 | 0 | 4.065 |
| CCL20 | 0 | 0 | 1.088 |
| CD6 | 0 | 0 | 1.959 |
| CDCP1 | 0 | 0.006 | 1.64 |
| CNTN2 | 0 | 0.002 | 3.128 |
| CXCL13 | 0 | 0 | 1.819 |
| CXCL9 | 0 | 0.005 | 1.419 |
| FLRT2 | 0 | 0 | 1.968 |
| GFAP | -0.05 | 0.005 | 1.692 |
| IL-12B | 0 | 0 | 2.062 |
| MOG | -0.04 | 0.003 | 1.267 |
| NfL | 0 | 0.008 | 0.5 |
| OPG | -0.014 | 0.002 | 2.743 |
| OPN | 0.045 | 0 | 4.172 |
| PRTG | 0 | 0 | 2.052 |
| SERPINA9 | 0 | 0 | 1.56 |
| TNFRSF10A | 0 | 0.003 | 0.565 |
| TNFSF13B | 0 | 0 | 3.75 |

Next, we tabulated the protein coefficients for each pathway. These were the *C*s in Eq. 4.

- Neuroinflammation:

- Immunomodulation:

Next, we tabulated the protein coefficients for each pathway. These were the Cs in Eq. 4. - Neuroinflammation:
| Feature | Coefficient |
| --- | --- |
| CCL20 | 0.397 |
| CD6 | 0.243 |
| CXCL13 | 0.668 |
| CXCL9 | 0.778 |
| IL-12B | -1.515 |
| TNFRSF10A | 0.089 |
| TNFSF13B | -2.705 |
| Intercept | -0.283 |

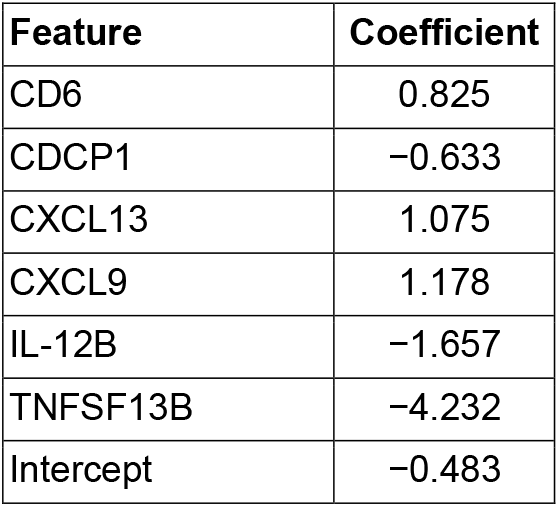
- Neuroaxonal Integrity:

- Immunomodulation:
| Feature | Coefficient |
| --- | --- |
| CD6 | 0.825 |
| CDCP1 | -0.633 |
| CXCL13 | 1.075 |
| CXCL9 | 1.178 |
| IL-12B | -1.657 |
| TNFSF13B | -4.232 |
| Intercept | -0.483 |

- Myelin Biology:

- Neuroaxonal Integrity:
| Feature | Coefficient |
| --- | --- |
| APLP1 | −0.149 |
| CNTN2 | −0.434 |
| FLRT2 | −0.321 |
| GFAP | 0.024 |
| NfL | 2.998 |
| OPG | 0.384 |
| OPN | 0.026 |
| PRTG | −0.621 |
| SERPINA9 | −0.779 |
| TNFRSF10A | −0.284 |
| Intercept | −0.744 |

- Myelin Biology:
| Feature | Coefficient |
| --- | --- |
| APLP1 | −1.206 |
| MOG | 2.653 |
| OPN | −0.037 |
| Intercept | −0.552 |

Then we tabulated the meta-classifier coefficients for the second stage of the stacked classifier. These were the *C*s in Eq. 3.

| Feature | Coefficient |
| --- | --- |
| APLP1 | −0.363 |
| CCL20 | 0.222 |
| CD6 | 0.119 |
| CDCP1 | −0.524 |
| CNTN2 | −0.21 |
| CXCL13 | 0.1 |
| CXCL9 | 0.252 |
| FLRT2 | −0.111 |
| GFAP | 0.042 |
| IL-12B | -0.432 |
| MOG | 0.485 |
| NfL | 2.038 |
| OPG | 0.056 |
| OPN | -0.072 |
| PRTG | 0.768 |
| SERPINA9 | 0.142 |
| TNFRSF10A | -0.178 |
| TNFSF13B | -1.3 |
| Neuroinflammation pathway classifier | 1.14 |
| Immunomodulation pathway classifier | 1.737 |
| Neuroaxonal integrity pathway classifier | 1.525 |
| Myelin biology pathway classifier | 0.435 |
| Intercept | -2.51 |

Finally, we tabulated the low and high values for scaling the probabilities in Eq.1 and Eq.2.

| **Disease Pathway/Disease Activity score** | **$P_{Low\ pth}$** | **$P_{High\ pth}$** |
| --- | --- | --- |
| **Disease Pathway** |  |  |
| Immunomodulation | 0.05 | 1.0 |
| Myelin Biology | 0.2 | 0.6 |
| Neuroaxonal Integrity | 0.1 | 1.0 |
| Neuroinflammation | 0.15 | 0.95 |
| **Disease Activity** | 0.05 | 1.00 |

Abbreviations: $P_{High\ pth}$ , high values for scaling probabilities; $P_{Low\ pth}$ , low values for scaling probabilities.

### FILE S2. Assay interferents

Samples were analyzed for potential interference with endogenous substances found naturally in patients’ samples (eg, lipemia, hemolysis, high protein count, and high bilirubin levels) and heterophilic antibodies such as HAMA and RF. Serum pools (*n*=4) were spiked with bilirubin, hemolysate, and lipids at typical concentrations using the ASSURANCE™ Interference Test Kit (Sun Diagnostics).

Heterophilic antibodies, including RF and HAMA, are established sources of potential interference in immunoassays. RF concentrate (Lee BioSolutions, Maryland Heights, MO, USA) was used to spike six serum samples at low (150 IU/mL) and high (2000 IU/mL) concentrations to determine the effect of RF interference on the analysis. For HAMA interference, five HAMA-positive serum samples with known established HAMA levels (ASSURANCE™ Interference Test Kit) were mixed at different ratios (10:90; 50:50; 90:10) with four MS samples from an internal cohort. Two HAMA-positive samples had a titer level >240 and three samples had a titer level >480, indicating that the positive samples had either >240 or >480 times more activity than a known negative. The mixed samples were compared with expected ratios of the individually measured samples to calculate the percent recovery.

Except for heterophilic antibody interference, the mean percentage recovery was calculated relative to the corresponding spike control (representing the same alteration of the serum sample without the addition of the interferent). For heterophilic antibody interference, the mean percentage recovery was calculated by comparing the RF spiked (low and high concentrations) samples with a corresponding spike control.

**TABLE S1.** Protein biomarkers included in assay panel to perform the MSDA Test

| **Biomarker** | **Full name** | **UniProt identifier** |
| --- | --- | --- |
| *APLP1* | Amyloid beta precursor-like protein 1 | P51693 |
| *CCL20* | C-C motif chemokine ligand 20 (MIP 3-alpha) | P78556 |
| *CD6* | Cluster of differentiation 6 | P30203 |
| *CDCP1* | CUB domain-containing protein 1 | Q9H5V8 |
| *CNTN2* | Contactin 2 | Q02246 |
| *CXCL13* | Chemokine (C-X-C motif) ligand 13 | P02462 |
| *CXCL9* | Chemokine (C-X-C motif) ligand 9 (MIG) | O43927 |
| *COL4A1*<sup>a</sup> | Collagen type IV alpha 1 | Q07325 |
| *FLRT2* | Fibronectin leucine-rich repeat transmembrane protein | O43155 |
| *GFAP* | Glial fibrillary acidic protein | P14136 |
| *GH*<sup>a</sup> | Growth hormone (somatotropin) | P01241 |
| *IL-12β* | Interleukin-12 subunit beta | P29460 |
| *MOG* | Myelin oligodendrocyte glycoprotein | Q16653 |
| *NfL* | Neurofilament light polypeptide chain | P07196 |
| *OPG* | Osteoprotegerin | O00300 |
| *OPN* | Osteopontin | P10451 |
| *PRTG* | Protogenin | Q2VWP7 |
| *SERPINA9* | Serpin family A member 9 | Q86WD7 |
| *TNFRSF10A* | Tumor necrosis factor receptor superfamily member 10A (TRAIL-R1) | O00220 |
| *TNFSF13B* | Tumor necrosis factor superfamily member 13B (BAFF) | Q9Y275 |
| *VCAN*<sup>a</sup> | Versican core protein | P13611 |

<sup>a</sup>These biomarkers were not used in the algorithm to determine Disease Pathway and Disease Activity scores.

**TABLE S2.**
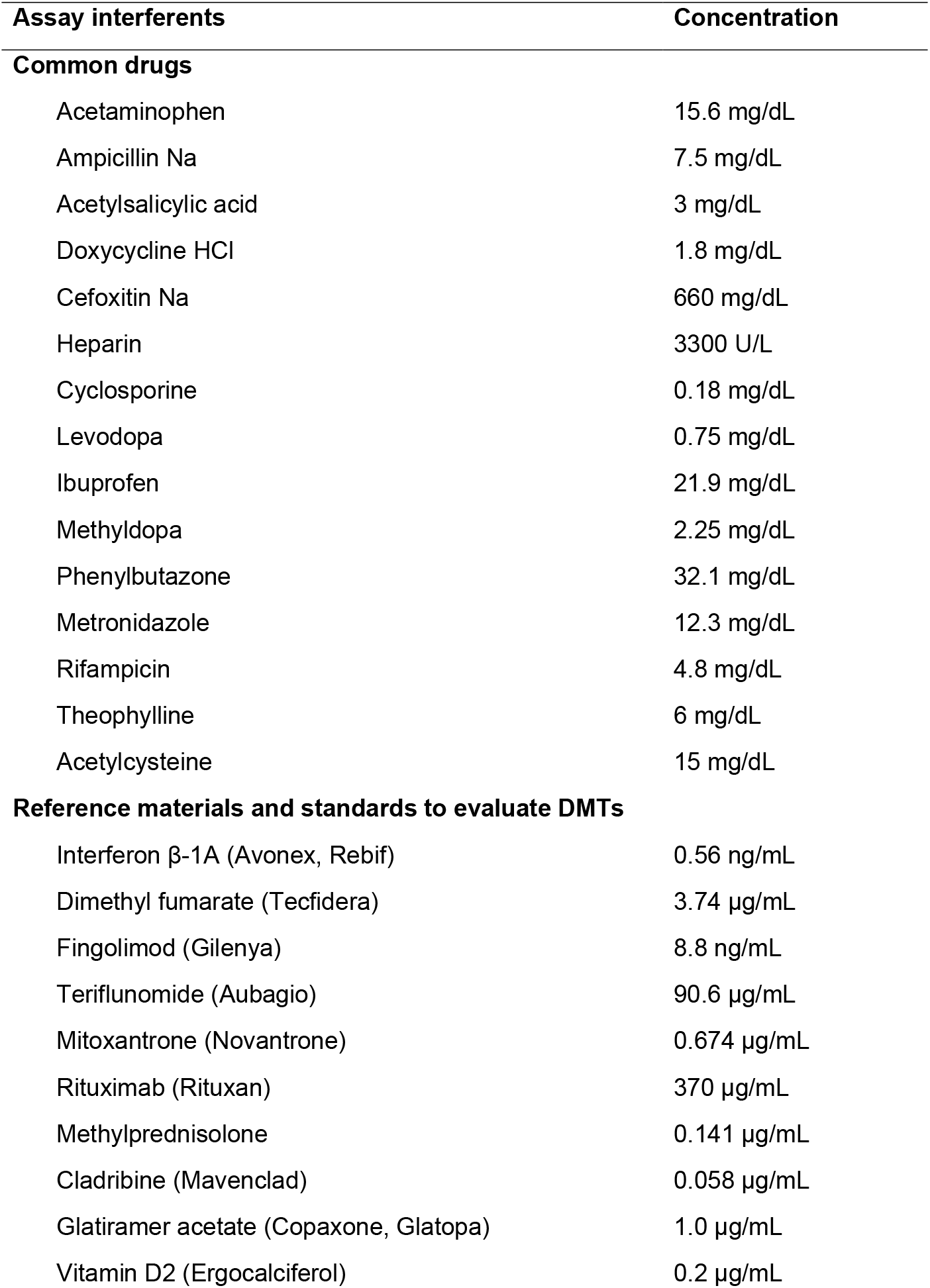
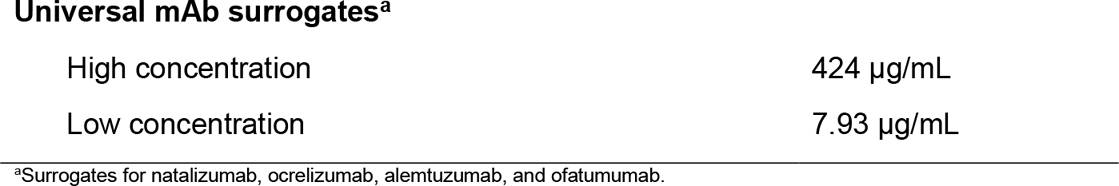
Common over-the-counter and prescription drugs, reference materials and standards, and mAbs evaluated for interference in the MSDA Test

**TABLE S3.** Lot to lot (batch 1 vs. batch 2) and laboratory to laboratory (development vs. clinical laboratory) consistency of the MSDA Test

|  | Mean % difference |  | R<sup>2</sup> correlation of samples |  |
| --- | --- | --- | --- | --- |
|  | Batch 1 vs. batch 2 | Development vs. clinical laboratory | Batch 1 vs. batch 2 | Development vs. clinical laboratory |
| *APLP1* | 0 | −1 | 0.93 | 0.95 |
| *CCL20* | −14 | −12 | 0.99 | 1.00 |
| *CD6* | −9 | −9 | 0.97 | 0.97 |
| *CDCP1* | −9 | −10 | 0.99 | 1.00 |
| *CNTN2* | 8 | 4 | 0.96 | 0.97 |
| *COL4A1* | −27 | −17 | 0.97 | 0.98 |
| *CXCL13* | −7 | −5 | 0.99 | 1.00 |
| *CXCL9* | −16 | −11 | 1.00 | 1.00 |
| *FLRT2* | −3 | −6 | 0.95 | 0.98 |
| *GFAP* | −13 | −9 | 0.96 | 0.97 |
| *GH* | −3 | −6 | 0.98 | 0.99 |
| *IL-12β* | −5 | −5 | 0.97 | 0.98 |
| *MOG* | 2 | 0 | 0.98 | 0.98 |
| *NfL* | −13 | −10 | 0.98 | 0.99 |
| *OPG* | −17 | −9 | 0.97 | 0.98 |
| *OPN* | −14 | −7 | 0.97 | 0.99 |
| *PRTG* | 0 | −5 | 0.97 | 0.98 |
| *SERPINA9* | −4 | −5 | 0.99 | 0.99 |
| *TNFRSF10A* | −7 | −11 | 0.98 | 0.99 |
| *TNFSF13B* | −6 | −5 | 0.96 | 0.98 |
| *VCAN* | −8 | −9 | 0.91 | 0.95 |

**TABLE S4.** Storage, processing, and stability of the individual biomarkers at various temperatures (room temperature, 4°C, and −20°C) over time during the initial and follow-up studies

| Mean % difference vs. experimental control conditions |  |  |  |  |  |  |  |  |  |  |  |  |  |  |  |  |  |  |  |  |  |
| --- | --- | --- | --- | --- | --- | --- | --- | --- | --- | --- | --- | --- | --- | --- | --- | --- | --- | --- | --- | --- | --- |
| Analyte | Initial study |  |  |  |  |  |  |  |  |  |  |  |  |  |  |  |  | Follow-up study |  |  |  |
|  | Room temperature |  |  |  |  |  | 4°C |  |  |  |  |  | -20°C |  |  |  |  | 4°C |  |  |  |
|  | 4 h | Day 1 | Day 3 | Day 7 | Day 14 | Day 28 | 4 h | Day 1 | Day 3 | Day 7 | Day 14 | Day 28 | Day 1 | Day 3 | Day 7 | Day 14 | Day 28 | Day 1 | Day 2 | Day 3 | Day 7 |
| APLP1 | 4 | 17 | 7 | 19 | 48 | 134 | -6 | -8 | 12 | 31 | 12 | 25 | 5 | 9 | 0 | 5 | 10 | 10 | 5 | 7 | 11 |
| CCL20 | 4 | 2 | -6 | -7 | 8 | 47 | 1 | -3 | 11 | 24 | 15 | 23 | 12 | 13 | 6 | 5 | 7 | -1 | -2 | 4 | 12 |
| CD6 | -2 | -4 | -14 | -5 | 19 | 93 | -10 | -11 | -7 | 7 | -2 | 10 | 1 | 5 | -1 | 1 | 1 | 2 | -1 | 2 | 5 |
| CDCP1 | -5 | -1 | -13 | -2 | 26 | 114 | -10 | -10 | -2 | 6 | 1 | 10 | 0 | 7 | -3 | 1 | 1 | 4 | 1 | 3 | 4 |
| CNTN2 | 0 | -3 | -13 | -3 | 24 | 71 | -8 | -10 | -9 | 2 | -3 | 7 | 2 | 6 | 1 | 2 | 3 | 3 | 0 | 1 | 1 |
| COL4A1 | -5 | -3 | -14 | -1 | 22 | 91 | -10 | -11 | -12 | 2 | -2 | 10 | 0 | 4 | 9 | 3 | 2 | 9 | -3 | 11 | 14 |
| CXCL13 | -1 | -7 | -25 | -27 | -24 | 7 | -5 | -6 | -8 | -6 | -4 | -3 | 6 | 12 | 4 | 7 | 5 | 2 | 0 | 0 | 1 |
| CXCL9 | -2 | -4 | -14 | -7 | 16 | 65 | -6 | -10 | -11 | -4 | -6 | -2 | 2 | 6 | 1 | 3 | 4 | 4 | 0 | 1 | 0 |
| FLRT2 | -4 | -1 | -11 | 0 | 26 | 97 | -10 | -10 | -2 | 11 | 2 | 13 | -1 | 8 | -1 | 1 | 2 | 3 | 0 | 2 | 3 |
| GFAP | 0 | 2 | -5 | 9 | 39 | 142 | -7 | -9 | -5 | 20 | 1 | 18 | 3 | 11 | -1 | 3 | 9 | 1 | 3 | 4 | 8 |
| GH | 1 | 0 | -10 | -6 | 9 | 52 | -6 | -9 | 2 | 10 | 7 | 12 | 4 | 9 | 1 | 5 | 7 | 2 | 3 | 2 | 10 |
| IL-12β | -2 | -13 | -34 | -40 | -32 | -24 | -7 | -11 | -14 | -18 | -9 | -15 | 3 | 8 | 0 | 3 | 4 | 3 | 0 | 1 | -4 |
| MOG | -2 | 0 | -10 | 1 | 23 | 83 | -7 | -9 | -1 | 13 | 2 | 12 | 1 | 6 | 1 | 3 | 3 | 1 | 2 | 1 | 3 |
| NfL | -3 | -6 | -11 | 2 | 29 | 110 | -11 | -9 | 1 | 4 | 3 | 12 | -4 | 4 | 0 | 2 | 2 | -3 | 6 | 1 | 6 |
| OPG | -2 | -4 | -13 | -5 | 18 | 94 | -6 | -9 | -8 | 1 | -1 | 5 | 1 | 6 | -1 | 2 | 2 | 2 | 0 | 1 | 1 |
| OPN | -4 | -5 | -18 | -13 | 1 | 47 | -6 | -16 | -27 | -24 | -26 | -26 | 0 | 5 | -1 | 3 | -1 | 2 | -2 | -2 | -8 |
| PRTG | -1 | -6 | -19 | -20 | -13 | -1 | -6 | -8 | -7 | -3 | -4 | -1 | 3 | 6 | 0 | 2 | 2 | 2 | 0 | 1 | 0 |
| SERPINA9 | -1 | -3 | -15 | -8 | 10 | 43 | -7 | -11 | -4 | 2 | 0 | 3 | 1 | 5 | -1 | 1 | 2 | 9 | 10 | 7 | 9 |
| TNFRSF10A | 0 | -2 | -11 | 0 | 31 | 108 | -7 | -11 | 1 | 11 | 3 | 10 | 0 | 9 | -2 | 0 | 2 | 5 | 0 | 2 | 5 |
| TNFSF13B | -4 | -7 | -20 | -10 | 19 | 108 | -10 | -14 | -12 | 0 | -6 | 1 | 1 | 7 | 1 | 3 | 2 | 3 | -1 | 3 | -1 |
| VCAN | -3 | -3 | -10 | -1 | 23 | 72 | -6 | -11 | -1 | 9 | 0 | 7 | 0 | 4 | -2 | 2 | 1 | 5 | 2 | 4 | 7 |

Green shading: %CV within $\pm 20\%$ . Red shading: %CV < -20% or >20%.

**TABLE S5.**
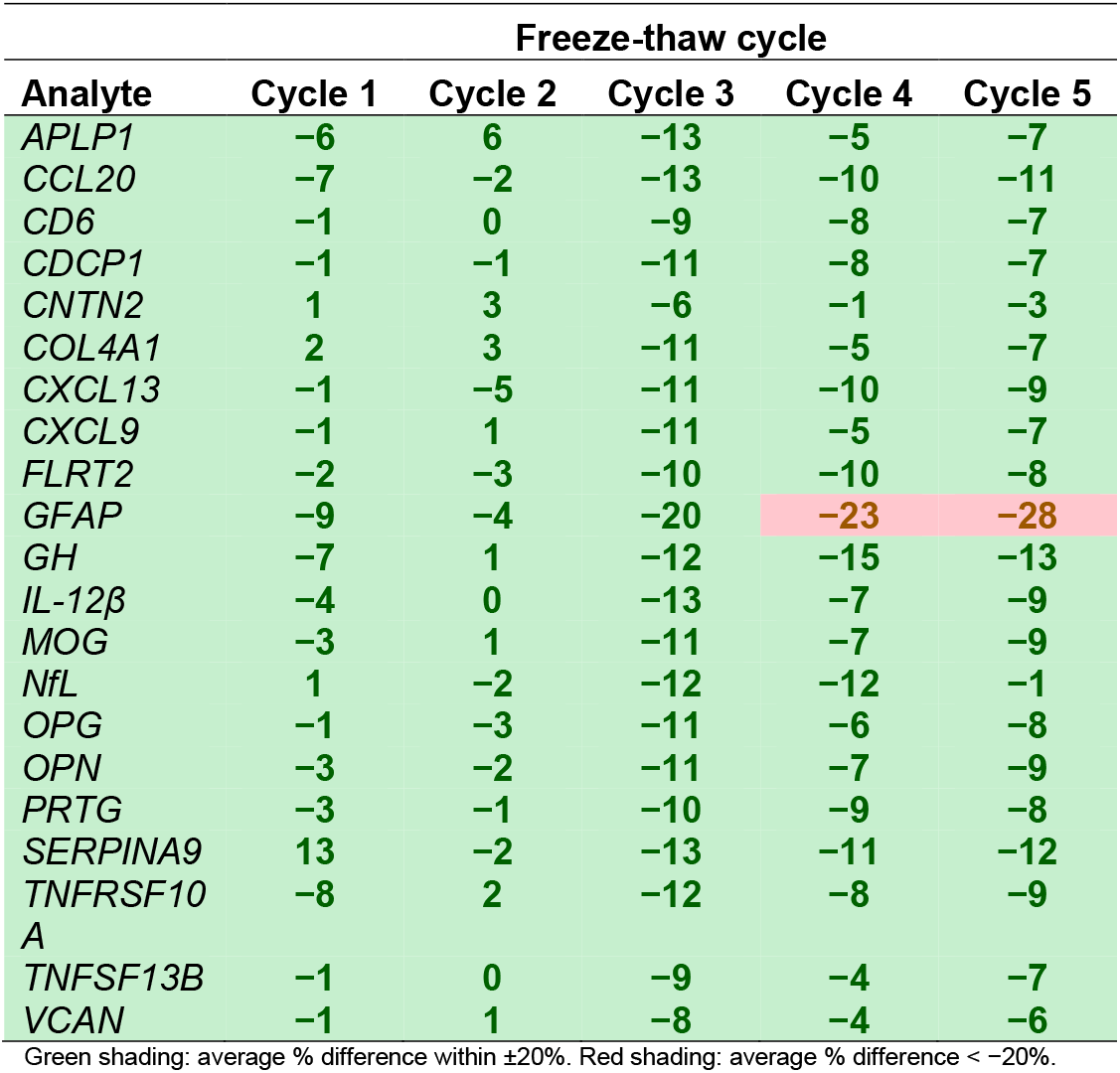
Storage, processing, and stability of the MSDA Test during various freeze-thaw cycles for individual biomarkers

| Analyte | Freeze-thaw cycle |  |  |  |  |
| --- | --- | --- | --- | --- | --- |
|  | Cycle 1 | Cycle 2 | Cycle 3 | Cycle 4 | Cycle 5 |
| APLP1 | -6 | 6 | -13 | -5 | -7 |
| CCL20 | -7 | -2 | -13 | -10 | -11 |
| CD6 | -1 | 0 | -9 | -8 | -7 |
| CDCP1 | -1 | -1 | -11 | -8 | -7 |
| CNTN2 | 1 | 3 | -6 | -1 | -3 |
| COL4A1 | 2 | 3 | -11 | -5 | -7 |
| CXCL13 | -1 | -5 | -11 | -10 | -9 |
| CXCL9 | -1 | 1 | -11 | -5 | -7 |
| FLRT2 | -2 | -3 | -10 | -10 | -8 |
| GFAP | -9 | -4 | -20 | -23 | -28 |
| GH | -7 | 1 | -12 | -15 | -13 |
| IL-12β | -4 | 0 | -13 | -7 | -9 |
| MOG | -3 | 1 | -11 | -7 | -9 |
| NfL | 1 | -2 | -12 | -12 | -1 |
| OPG | -1 | -3 | -11 | -6 | -8 |
| OPN | -3 | -2 | -11 | -7 | -9 |
| PRTG | -3 | -1 | -10 | -9 | -8 |
| SERPINA9 | 13 | -2 | -13 | -11 | -12 |
| TNFRSF10A | -8 | 2 | -12 | -8 | -9 |
| TNFSF13B | -1 | 0 | -9 | -4 | -7 |
| VCAN | -1 | 1 | -8 | -4 | -6 |

Green shading: average % difference within $\pm 20\%$ . Red shading: average % difference $< -20\%$ .

**TABLE S6.**
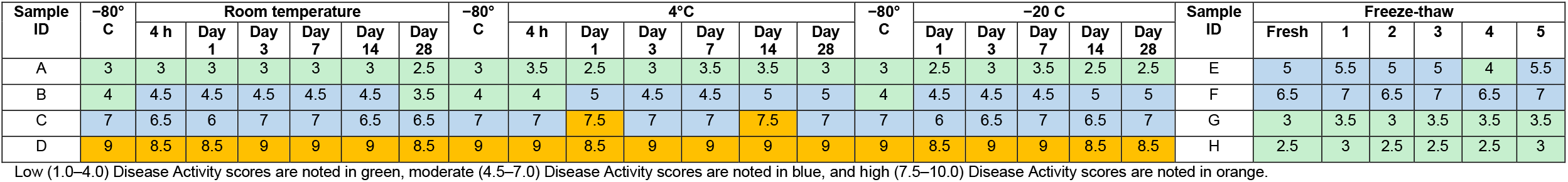
Initial study of the storage, processing, and stability of the MSDA Test at various temperatures (room temperature, 4°C, and −20°C) over time and during freeze-thaw at the Disease Activity score level

| Sample ID | –80° C | Room temperature |  |  |  |  |  | –80° C | 4°C |  |  |  |  |  | –80° C | –20 C |  |  |  |  | Sample ID | Freeze-thaw |  |  |  |  |  |
| --- | --- | --- | --- | --- | --- | --- | --- | --- | --- | --- | --- | --- | --- | --- | --- | --- | --- | --- | --- | --- | --- | --- | --- | --- | --- | --- | --- |
|  |  | 4 h | Day 1 | Day 3 | Day 7 | Day 14 | Day 28 |  | 4 h | Day 1 | Day 3 | Day 7 | Day 14 | Day 28 |  | Day 1 | Day 3 | Day 7 | Day 14 | Day 28 |  | Fresh | 1 | 2 | 3 | 4 | 5 |
| A | 3 | 3 | 3 | 3 | 3 | 3 | 2.5 | 3 | 3.5 | 2.5 | 3 | 3.5 | 3.5 | 3 | 3 | 2.5 | 3 | 3.5 | 2.5 | 2.5 | E | 5 | 5.5 | 5 | 5 | 4 | 5.5 |
| B | 4 | 4.5 | 4.5 | 4.5 | 4.5 | 4.5 | 3.5 | 4 | 4 | 5 | 4.5 | 4.5 | 5 | 5 | 4 | 4.5 | 4.5 | 4.5 | 5 | 5 | F | 6.5 | 7 | 6.5 | 7 | 6.5 | 7 |
| C | 7 | 6.5 | 6 | 7 | 7 | 6.5 | 6.5 | 7 | 7 | 7.5 | 7 | 7 | 7.5 | 7 | 7 | 6 | 6.5 | 7 | 6.5 | 7 | G | 3 | 3.5 | 3 | 3.5 | 3.5 | 3.5 |
| D | 9 | 8.5 | 8.5 | 9 | 9 | 9 | 8.5 | 9 | 9 | 8.5 | 9 | 9 | 9 | 9 | 9 | 8.5 | 9 | 9 | 8.5 | 8.5 | H | 2.5 | 3 | 2.5 | 2.5 | 2.5 | 3 |

Low (1.0–4.0) Disease Activity scores are noted in green, moderate (4.5–7.0) Disease Activity scores are noted in blue, and high (7.5–10.0) Disease Activity scores are noted in orange.

**TABLE S7.** Follow-up study of the storage, processing, and stability of the MSDA Test at 4°C over time at the Disease Activity score level

| Sample ID | −80°C | 4°C |  |  |  |
| --- | --- | --- | --- | --- | --- |
|  |  | Day 1 | Day 2 | Day 3 | Day 7 |
| A | 8.5 | 8.5 | 8.5 | 8.5 | 8.5 |
| B | 5 | 4.5 | 4.5 | 4.5 | 4.5 |
| C | 7.5 | 6 | 7.5 | 7.5 | 7 |
| D | 6.5 | 6.5 | 6.5 | 5.5 | 7 |
| E | 5 | 5 | 5 | 5 | 5.5 |
| F | 7 | 7 | 7 | 6.5 | 7.5 |
| G | 5.5 | 5 | 5 | 5 | 5.5 |
| H | 6.5 | 6.5 | 7.5 | 7.5 | 7.5 |
| I | 6 | 6 | 6.5 | 6 | 7 |
| J | 3.5 | 3.5 | 3 | 3.5 | 3 |
| K | 1.5 | 1 | 1.5 | 1.5 | 1.5 |
| L | 2.5 | 2.5 | 2.5 | 2.5 | 2 |
| M | 2.5 | 2.5 | 3 | 2 | 3 |
| N | 7.5 | 7.5 | 8.5 | 8 | 8 |

Low (1.0–4.0) Disease Activity scores are noted in green, moderate (4.5–7.0) Disease Activity scores are noted in blue, and
high (7.5–10.0) Disease Activity scores are noted in orange.

**FIGURE S1.**
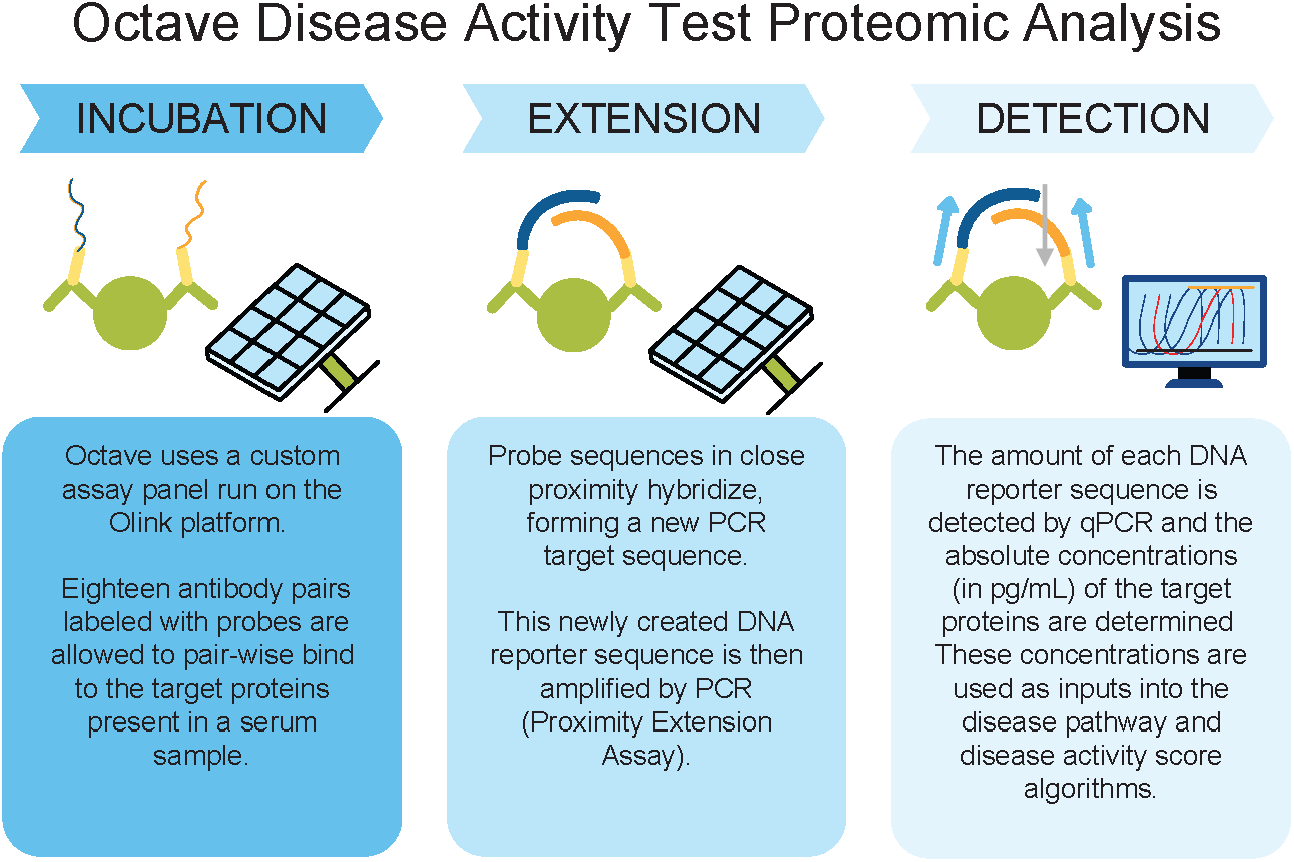
Overview of PEA technology and MSDA Test format. Eighteen antibody pairs, labeled with DNA oligonucleotides, bind to target antigen in solution. Oligonucleotides that are brought into proximity will hybridize and are extended by a DNA polymerase. This newly created piece of DNA barcode is amplified by PCR. The amount of each DNA barcode is quantified by microfluidic qPCR (Biomark™ HD, Fluidigm, Maryland Heights, MO, USA), with results reported in cycle threshold values. Data processing is then performed in the Olink^®^ NPX Manager (Olink Proteomics, Uppsala, Sweden) software to convert the Ct values to a Normalized Protein eXpression value. The signal obtained from the assay (Normalized Protein eXpression) is converted to absolute concentration (pg/mL) using three calibrators that cover the range of sample response in the MS population (calibrators: high, middle, low) and then referenced back to the standard curve. These concentrations are then used as inputs into algorithms corresponding to disease activity and biological pathway scores.

**FIGURE S2.**
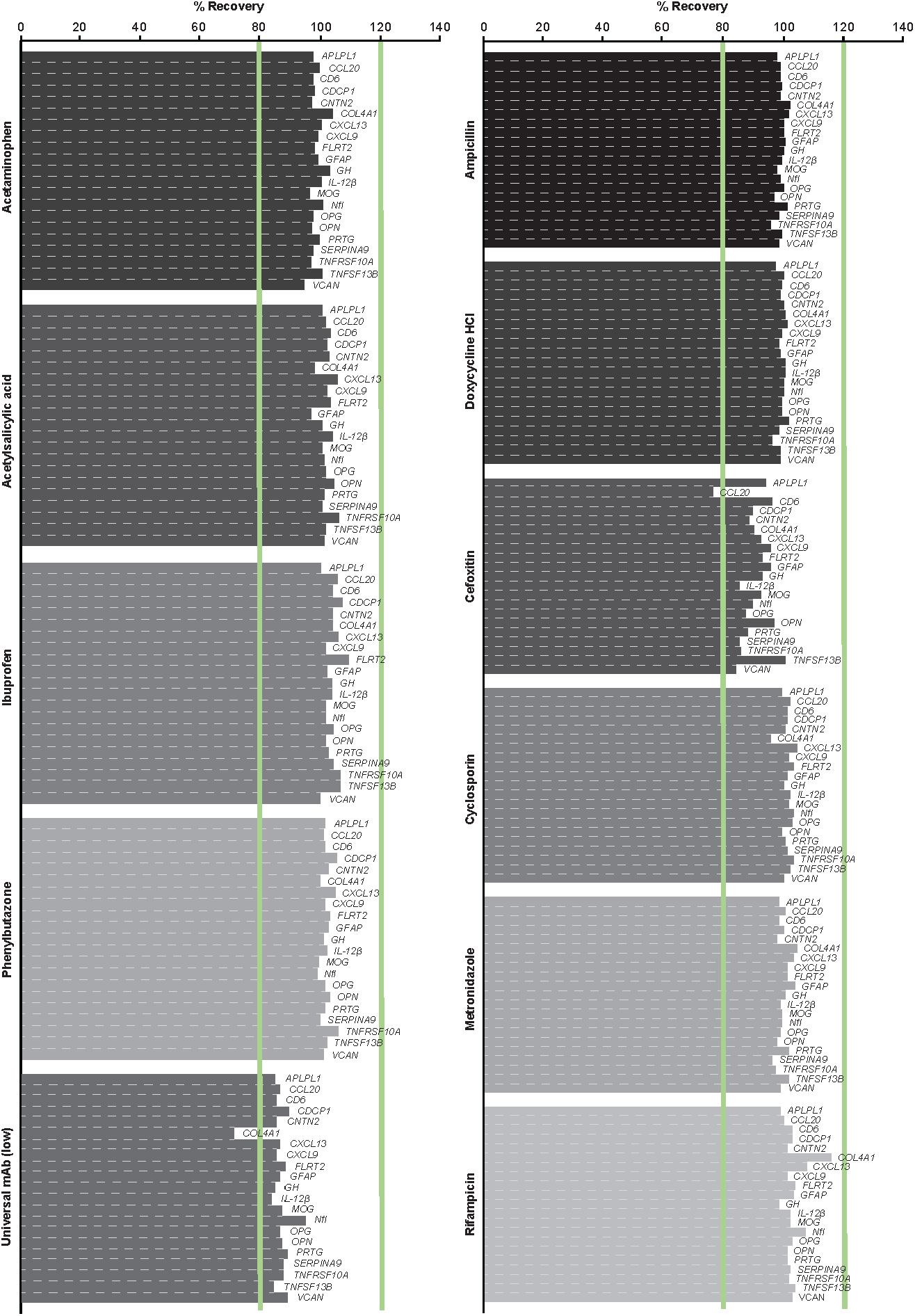
Assay interference using common drugs.

**FIGURE S3.**
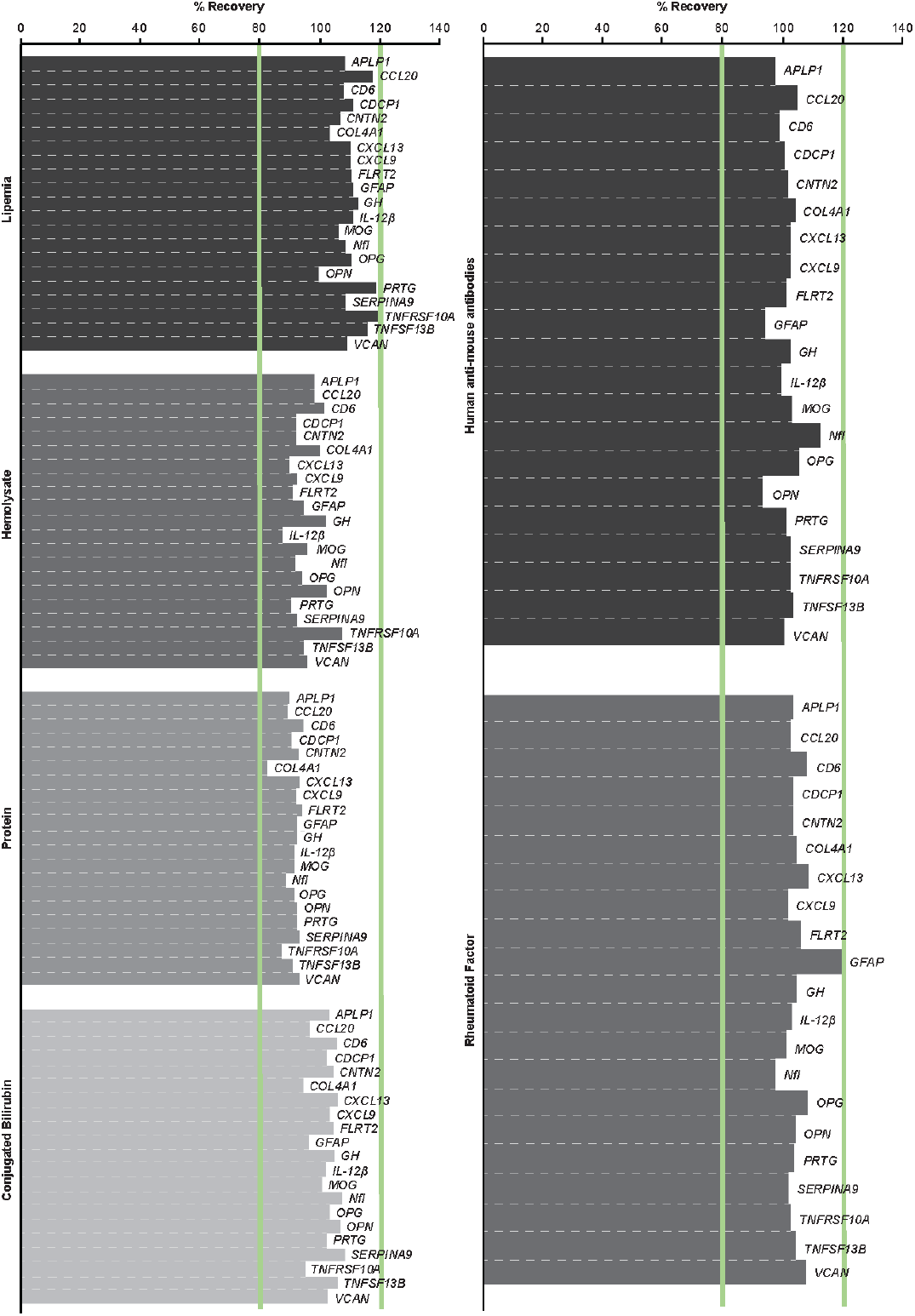
Assay interference using routine endogenous interferents and heterophilic antibodies.

**FIGURE S4.**
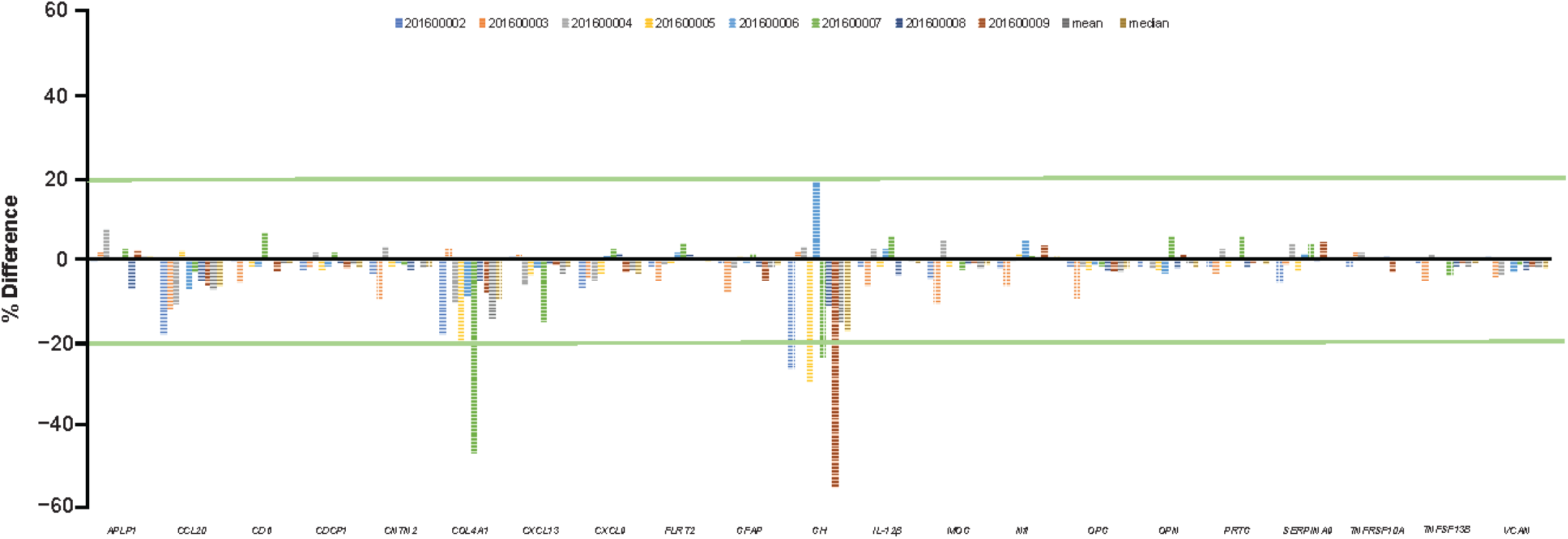
Percent difference of the observed protein concentration relative to the average concentration as determined from six time points (days 1, 2, 3, 4, 5, and 12) in eight samples assayed in the MSDA Test.

## Notes

### Author Declarations

The study used both publicly available data and data acquired under institutional agreement. Serum samples from publicly available sources were analyzed for analytical validation experiments. Serum samples from patients with multiple sclerosis (MS) were analyzed during the assay development and validation process to establish the MS reference ranges for each analyte. These samples came from the following institutional sources: Accelerated Cure Project (Waltham, MA, USA), Brigham and Women's Hospital (Boston, MA, USA), University of California San Francisco (San Francisco, CA, USA), University Hospital Basel (Basel, Switzerland), American University Beirut (Beirut, Lebanon), University of Massachusetts (Boston, MA, USA), University of Pittsburgh (Pittsburgh, PA, USA) and Rocky Mountain Multiple Sclerosis Clinic (Salt Lake City, UT, USA). Data were acquired under institutional agreement and the study was approved by each institutional review board. All patients provided written informed consent.

### Summary of Updates

The MS Disease Activity (MSDA) Test is a multi-protein, serum-based biomarker assay designed to quantitatively measure disease activity using the protein levels of biomarkers present in the serum of patients with MS. In this study, we evaluated 21 biomarkers, 18 of which were selected for inclusion in the MSDA Test, and extensively characterized the MSDA Test (individual biomarkers and algorithmic scores) by establishing the accuracy, precision, sensitivity, and robustness of the assay. This study serves as a critical first step in the validation of this multi-protein, serum-based assay, which will be a quantitative, minimally invasive, and scalable tool to improve MS disease management.

